# Microstructural degeneration underlies executive dysfunction after stroke

**DOI:** 10.1101/2020.02.03.20020131

**Authors:** Michele Veldsman, Emilio Werden, Natalia Egorova, Mohamed Salah Khlif, Amy Brodtmann

**Affiliations:** Department of Experimental Psychology, University of Oxford, Oxford, UK; The Florey Institute of Neuroscience and Mental Health, University of Melbourne, Melbourne, Australia; Melbourne School of Psychological Sciences, University of Melbourne, Melbourne, Victoria, Australia; Austin Health, Heidelberg, Melbourne, Victoria, Australia; Eastern Clinical Research Unit, Box Hill Hospital, Melbourne, Victoria, Australia

## Abstract

**Objective:** Executive dysfunction affects 40% of stroke patients and is associated with poor quality of life. Stroke severity and lesion volume rarely predict whether a patient will have executive dysfunction. Stroke typically occurs on a background of cerebrovascular burden, which impacts cognition and brain network structural integrity. We investigated whether measures of white matter microstructural integrity and cerebrovascular risk factors better explain executive dysfunction than markers of stroke severity.

**Methods:** We used structural equation modelling to examine multivariate relationships between cerebrovascular risk, white matter microstructural integrity (fractional anisotropy and mean diffusivity), stroke characteristics and executive dysfunction in 126 stroke patients (mean age 68.4 years), three months post-stroke, and compared to 40 age- and sex-matched control participants. Executive function was measured using the Trail Making Tests, Clock Drawing task and Rey Complex Figure copy task. Microstructural integrity was estimated using a standard pipeline to process diffusion weighted images.

**Results:** Executive function was below what would be expected for age and education level in stroke patients (t-test compared to controls t(79)=5.75, p<0.001). A multivariate structural equation model illustrated the complex relationship between executive function, white matter integrity, stroke characteristics and cerebrovascular risk. Pearson’s correlations confirmed a stronger relationship between executive dysfunction and white matter integrity, than executive dysfunction and stroke severity. Mediation analysis showed the relationship between executive function and white matter integrity is mediated by cerebrovascular burden.

**Interpretation:** White matter microstructural degeneration of the superior longitudinal fasciculus in the executive control network better explains executive dysfunction than markers of stroke severity.

## INTRODUCTION

Executive function is impaired in around 40% of ischemic stroke patients^1^ and predicts quality of life, after controlling for age, depression and stroke severity^2^. It is not currently possible to predict executive dysfunction based on characteristics of a patient’s stroke, such as location or severity, alone. Despite relatively focal damage caused by stroke, remote effects occur across distributed brain networks^3^. Structural and functional brain network integrity is critical to maintaining cognitive function^4^.

Brain networks show a high degree of functional specificity^5^. The canonical frontoparietal brain network subserves higher cognitive functions related to attention and executive function^6,7^. The superior longitudinal fasciculus (SLF) is the principal white matter tract connecting the frontal and parietal nodes of the frontoparietal network^8^. Within the SLF, dorsal, middle and ventral branches connect superior, intraparietal and inferior parietal and frontal regions to superior, middle and inferior frontal regions, respectively^8^: an architecture preserved from the monkey brain.

Collectively, these divisions facilitate communication between regions of the frontoparietal network underlying broad executive control functions^6,8^. Although no agreed definition of executive function exists, most investigators agree higher order cognitive components of executive function include planning and maintaining strategies for controlling and executing behavior, allocating attention, and inhibiting distracting information^9^.

There has been no systematic investigation relating stroke location to executive impairment, likely because stroke imaging cohorts have been too small and variable to detect consistent relationships between different cognitive functions and stroke locations. In addition, stroke has widespread effects across the brain^3,10^. There is some evidence that strokes affecting anterior circulation^9^ or frontal circuits^11^ are more likely to result in executive impairment, but a number of authors found executive impairment in cohorts of stroke patients with a mix of stroke locations and etiologies.^12–15^

Stroke occurs on a background of aging and cerebrovascular risk (CVR) factors which predominantly impact executive function^16,17^. Modifiable risk factors that contribute to cerebrovascular burden, such as smoking and hypertension, impact brain network integrity^18,19^. The best known macroscale markers of cerebrovascular burden are white matter hyperintensities (WMHs)^20^. Yet the relationship between WMHs and executive function is far from clear^21^. Vascular risk factors may also impact on the *normal appearing* white matter of brain networks^22^. Microstructural markers measured from magnetic resonance diffusion weighted images can act as surrogate markers of brain network integrity. These markers show a decline in ageing that is accelerated in the presence of CVR factors. For example, high blood pressure is associated with disrupted white matter microstructural integrity, even when controlling for age and the presence of white matter hyperintensities^23^. Decreased executive function was associated with altered functional connectivity of the frontoparietal network in 1007 elders, with reduced white matter integrity specific to the SLF in hypertensive compared to normotensive individuals^24^.

A multifaceted model likely explains executive function after stroke. We hypothesized that executive dysfunction would be well predicted by the integrity of the superior longitudinal fasciculus, the major associative tract in the frontoparietal network that is sensitive to cerebrovascular burden. We posited that this relationship would be mediated by vascular risk burden.

## METHODS

We analyzed data from 135 stroke patients and 40 healthy controls from the Cognition and Neocortical Volume After Stroke study (CANVAS)^25^, a hospital ethics committee approved study. All participants gave written, informed consent. Patients with hemorrhagic stroke, significant comorbidities, pre-existing dementia, or without a proven clinical ischemic stroke on imaging were excluded from participation.

Patients underwent MRI scanning and comprehensive neuropsychological testing at repeated intervals over three years. Healthy, age matched controls underwent the same sessions at equivalent intervals. The data from three months post-stroke were analysed here. Participants were interviewed to record vascular risk factors and medications, with information cross-referenced against hospital records. As part of a battery of neuropsychological testing, participants completed the Trail-Making Test (B)^26^, the Clock Drawing Task^27^ and the Rey Complex Figure Test^28^. These tests were included in the executive function domain score.

### Cerebrovascular risk score

A cerebrovascular risk score was calculated for each participant based on the sum of known risk factors. Individual risk factors were thresholded according to published guidelines for the management of cerebrovascular risk^29^. A body mass index greater than 30kg/m2, diagnosed or medicated hypertension, cholesterol, Type II Diabetes, ischemic heart disease, or atrial fibrillation contributed to the score. Positive smoking status and drinking greater than 14 units per week also contributed to the score. We used a bespoke score created with the available vascular risk data collected as part of the CANVAS^25^ study.

### Image acquisition

Magnetic resonance images were acquired on a Siemens 3T scanner with a 12-channel head coil. MRI included a fluid attenuated inversion recovery image (FLAIR), diffusion weighted imaging (DWI) and high-resolution structural magnetization prepared rapid acquisition gradient echo (MPRAGE) image, as part of a longer protocol^25^. A 160 sagittal slice MPRAGE had 1mm isotropic voxels, 1900ms repetition time (TR), 2.55ms echo time (TE), 9°flip angle and 256×256 acquisition matrix. The 3-D SPACE FLAIR had 160, 1mm thick sagittal slices, a TR of 6000ms, a TE of 380ms, a 120° flip angle and a 256×254 acquisition matrix. Sixty volumes of single shot spin echo EPI with a TR 8.4s, TE 110ms were collected, with 60 diffusion sensitization directions, *b*=3000 s/mm^2^ and 2.5mm isotropic voxels. Additional *b*=0 reverse phase-encoded images were acquired to aid in geometric distortion correction.

### Imaging analysis

Stroke infarcts were verified by a stroke neurologist (AB) and manually delineated by an imaging researcher expert in lesion tracing (MSK). The total volume of lesion was estimated from the binary manual mask. White matter hyperintensities (WMH) were segmented automatically using the Wisconsin WMH Segmentation Toolbox (W2MHS)^30^. Classification of WMHs was based on a supervised Random Forest-based regression. Both 3D FLAIR and MPRAGE images were used in the segmentation process. Total WMH load was calculated for each participant.

Diffusion images were preprocessed in the Diffusion Toolbox (FDT) in the fMRIB Software Library (FSL) using a standard pipeline. The “topup” function in FDT estimated susceptibility induced distortions and was input into the “eddy” function for correction of movement and gradient coil distortions. Diffusion tensors were fit using the inbuilt DTIFIT function. Fractional anisotropy (FA) and mean diffusivity (MD) values were extracted from the superior longitudinal fasciculus (SLF) as defined by the John Hopkins University tractography atlas^31^. Nine stroke participants were excluded due to imaging artefacts in their scans as a result of movement or problems with registration of images that could not be manually fixed.

### Statistical analysis

Executive function was compared in the stroke and healthy control group using Welch’s independent t-tests, Holm-Bonferroni corrected for multiple comparisons, in the JASP statistical package. Performance in each test was normalized against age-matched, published norms. Multiple regression was used to further explore this relationship correcting for age and years of education between groups.

Structural equation modelling (SEM) was carried out in the R Lavaan Package ^32^. Only stroke patients were included in the SEM as central to the model was the impact of stroke characteristics. The measurement model included a latent variable representing frontoparietal white matter integrity, made up of mean FA and MD of the superior longitudinal fasciculus. A second latent variable represented executive impairment with indicators based on performance in the three executive cognitive tasks. A third latent variable indexed stroke severity and included the volume of the lesion, the National Institute of Health Stroke Scale (NIHSS) score and the modified Rankin Scale (mRS) three months post-stroke. Confirmatory factor analysis predicted the contribution of the latent variables to their indicator variables. Observed variables included age, WMH load and cerebrovascular risk score. Missing data were estimated using full information maximum likelihood (FIML). Nested model testing was used to confirm the model was a good fit for the data and assess the contribution of the frontoparietal white matter integrity latent variable. Multiple model fit indices were used to determine if the model was a good fit for the data.

Latent variables were extracted using a built-in function in the Lavaan package and correlated with observed variables using Pearson’s correlations and a significance level set to p<0.05. Mediation analysis was used to further clarify the relationship between executive dysfunction, frontoparietal white matter integrity, stroke severity and cerebrovascular risk.

## Data availability

The data that support the findings of this study are available on reasonable request from the corresponding author. The data are not publicly available as CANVAS is a prospective, “live” study, with expected completion of data acquisition in mid-2020 for the 5-year scanning timepoint. All requests for raw and analysed data will be reviewed by the CANVAS investigators to determine whether the request is subject to any intellectual property or confidentiality obligations.

## Results

Stroke patients and controls were well matched in terms of age and ratio of male to female (Table 1). As can be seen in the stroke lesion overlap map (Figure 1), the sites of stroke damage varied across patients with more right than left hemisphere strokes.

**Table 1:** Patient and control demographics Group demographics at three month timepoint. Abbreviations: IQR-interquartile range, SD-standard deviation, MoCA – Montreal Cognitive Assessment, CVR – cerebrovascular risk score, NIHSS – National Institute of Health Stroke Scale, mRS -Modified Rankin Scale

| Demographics | Statistics |  | Group Difference |
| --- | --- | --- | --- |
|  | Stroke patients | Healthy Controls |  |
| Number (female) | 135(41) | 40(15) | Chi Square $X^2=0.72$,<br>p=0.40 |
| Mean Age (SD) | 68 (11.8) | 69 (6.6) | Welch's t(120)=0.26,<br>p=0.80 |
| Median years of education<br>(IQR 1,3) | 12<br>(10,15) | 17<br>(11,18) | Mann-Whitney w=3689,<br>p<0.001 |
| Mean MoCA (SD) | 23 (4.1) | 26 (2.2) | Welch's t(119)=4.99,<br>p<0.001 |
| Median NIHSS (IQR 1,3) | 0 (0,1) | - | - |
| Median mRS (IQR 1,3) | 1 (1,2) | - | - |
| Median CVR Score (IQR 1,3) | 2 (2,4) | 1 (1,2) | Mann-Whitney w=-<br>1712, p<0.001 |
| Body Mass Index (%<br>>30kg/m<sup>2</sup>) | 30 | 18 | - |
| Percent diagnosed/medicated<br>hypertension | 63 | 43 | - |
| Percent diagnosed/medicated<br>hypercholesteremia | 46 | 35 | - |
| Percent with Type II Diabetes | 34 | 10 | - |
| Percent with Ischaemic heart<br>disease | 25 | 0.03 | - |
| Percent diagnosed atrial<br>fibrillation | 24 | 0.03 | - |
| Percentage of smokers | 47 | 33 | - |
| Percent drinking > 14 units<br>alcohol per week | 10 | 18 | - |
| Mean stroke lesion volume in<br>mm<sup>3</sup> (SD) | 7979<br>(13810) | - | - |
| Mean white matter<br>hyperintensity volume mm<sup>3</sup> (SD) | 19 (8.3) | 10 (2.7) | Welch's t(145)=-9.78,<br>p<0.0001 |
| Mean fractional anisotropy in superior longitudinal fasciculus (SD) | 0.43<br>(0.04) | 0.46<br>(0.01) | Welch's t(144)=6.52,<br>p<0.0001 |
| Mean mean diffusivity in the superior longitudinal fasciculus (cube transformed)(SD) | 0.08<br>(0.002) | 0.08<br>(0.00001) | Welch's t(134)=-5.52,<br>p<0.0001 |

**Figure 1:**
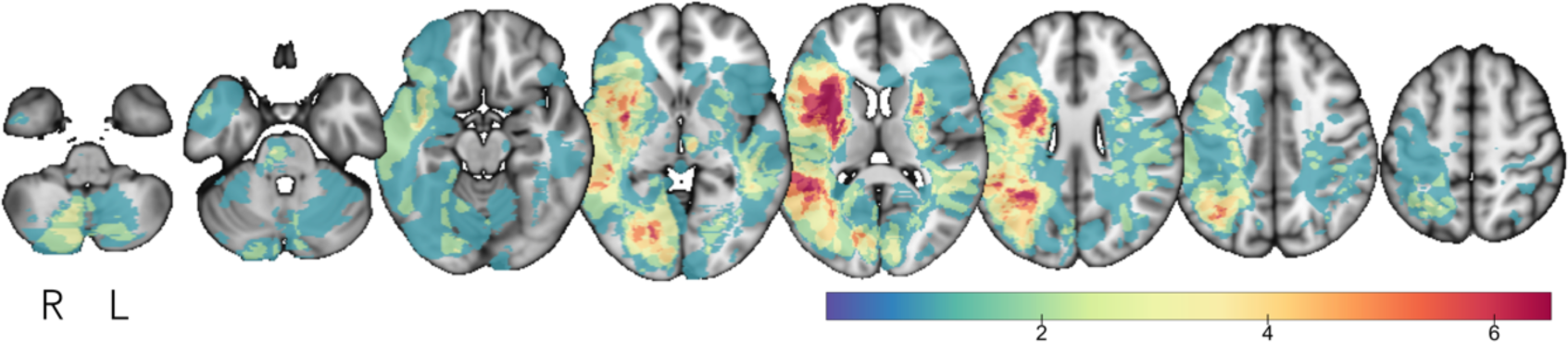
Lesion overlap map. Lesion overlap map displayed on Montreal Neurological Institute template. Colorbar indicates maximum number of overlapping lesions.

**Figure 2:**
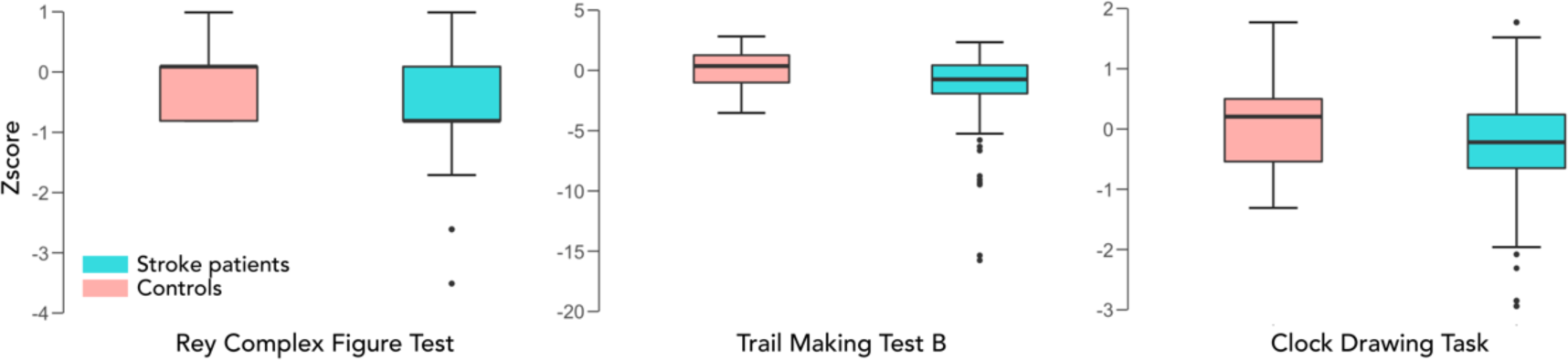
Executive dysfunction in stroke patients compared to age-matched controls. Boxplots displaying performance (z-score) in stroke patients compared to age-matched controls in three tests of executive function, the Rey Complex Figure task (left panel), Trail Making Test B (middle panel), Clock Drawing task (right panel).

Welch’s independent t-tests confirmed that stroke patients were impaired on the Rey Complex Figure Test, t(78)=5.75, p<0.001, and the Trail-Making Test (B), t(125)=4.23, p<0.001. Patients and healthy controls performed to a similar level on the Clock Drawing Task t(82)=1.81, p=0.07. Given a significant difference in the median years of education between stroke patients and healthy controls (Table 1), multiple regressions were run to control for age and education level. Even after controlling for age and education level, stroke was a significant predictor of performance in the Rey Complex Figure Test, F(3,155)=17, R^2^=0.25, p<0.001 (group significant predictor at p<0.001); and the Trail-Making Test (B), F(3,157)=3.70, R^2^=0.07, p<0.01 (group significant predictor at p<0.001) but not the Clock Drawing Task, F(3,160)=2.01, R^2^=0.04, p=0.12 (group non-significant predictor at p=0.13).

Multiple model fit indices confirmed that our full structural equation model (Figure 3a) was a very good fit for the data. Comparative Fit index was 0.99 for the full model (Figure 3a) and 0.80 for the constrained model (Figure 3b). The Tucker-Lewis Index, which penalizes overly complex models, was 0.99 for model A and 0.71 for model B. For both these fit indices, values greater than 0.9 indicate good model fit^33^. The absolute fit index, root mean square error of approximation (RMSEA), was 0.02 for model a and 0.10 for model b. As a guideline, RMSEA less than 0.05 is considered a very good fit for the data^33^. Analysis of variance in Lavaan was used to compare nested model fits and confirmed the full model (model a) was superior to model b, p<0.0001.

**Figure 3:**
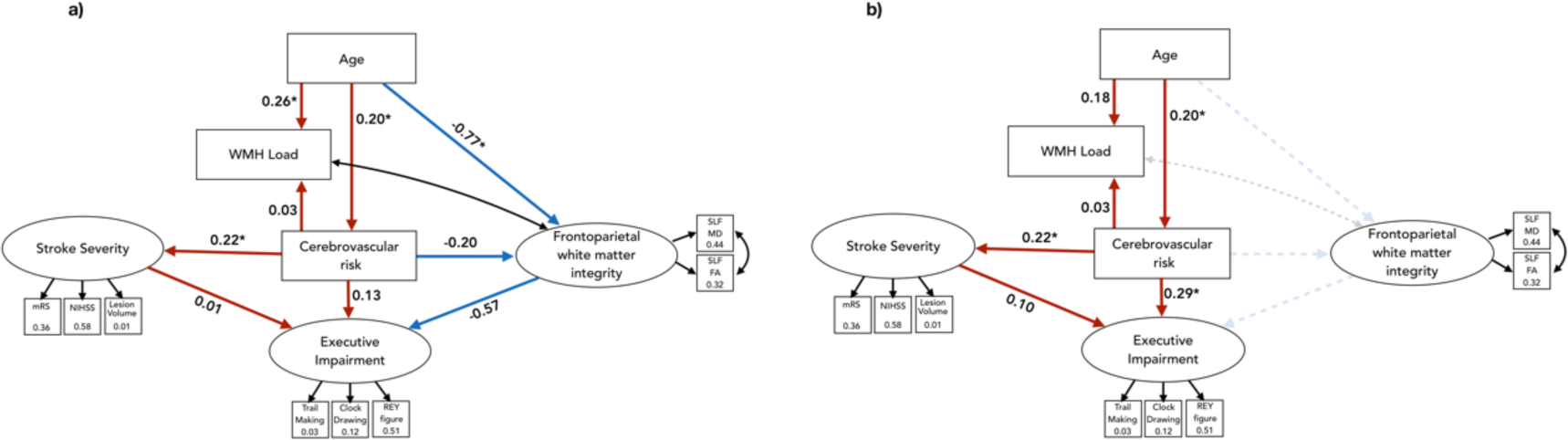
Structural equation path models. Values in square indicators are R^2^. Values on paths are standardized beta estimates. a) Full path model. b) Model constrains the contribution of frontoparietal white matter integrity of the superior longitudinal fasciculus for nested model testing. Red paths show positive directional relationships, blue paths show negative relationships. Black arrows indicate covariances. *paths significant to p<0.05

There was a significant Pearson’s correlation between the latent variable representing executive function and stroke severity r=0.22, p<0.01, and executive function and white matter integrity r=-0.74, p<0.001 but not between executive function and cerebrovascular burden, r=0.15, p=0.09. Pearson and Filon’s z test to compare the magnitude of overlapping correlations confirmed that the magnitude of the correlation between executive function and frontoparietal white matter integrity was significantly greater than the magnitude of the correlation between executive function and stroke severity, z(123)=-9.68, p<0.001 (Fig 4). Both measures of white natter integrity used in the latent variable (fractional anisotropy and mean diffusivity); median cerebrovascular risk score and white matter hyperintensity load were all significantly different in stroke patients compared to age-matched controls (Table 1). This confirms reduced white matter integrity of the SLF, increased white matter hyperintensity and higher cerebrovascular risk burden in stroke patients compared to age-matched controls.

**Figure 4:**
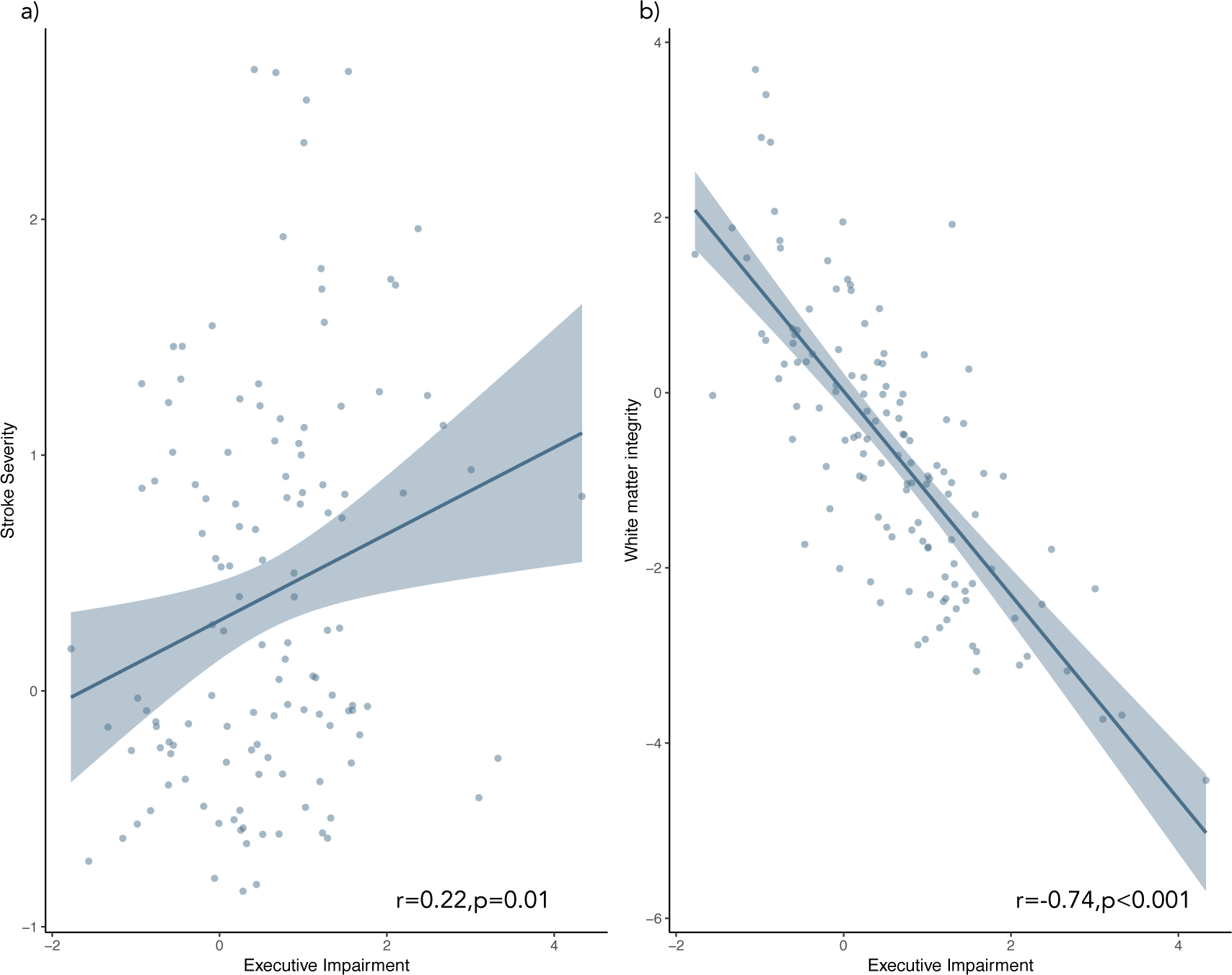
Scatterplots showing latent variable relationships with executive impairment. Correlation between executive impairment and a) a latent variable representing stroke severity and b) a latent variable representing white matter integrity of the superior longitudinal fasciculus

Mediation analysis was used to assess the contribution of cerebrovascular risk (Figure 5a) and stroke severity (Figure 5b) to the relationship between frontoparietal white matter integrity and executive impairment. Both models were a very good fit for the data (CFI and TLI =1, RMSEA <0.001 for both models). Confirming the results of the SEM, executive impairment was associated with frontoparietal white matter integrity. Both cerebrovascular risk and stroke severity predicted frontoparietal white matter integrity (standardized beta estimates -0.20 and -0.45, respectively, p<0.001). In both models, when the relationship between frontoparietal white matter integrity and executive function is mediated by either CVR or stroke severity the relationship became non-significant (standardized beta estimates -0.003 and -0.02 respectively) indicating complete mediation (Figure 5). Cerebrovascular risk (CVR) reduced the relationship between frontoparietal white matter integrity and executive impairment to a greater extent than stroke severity suggesting it has a stronger mediating effect.

**Figure 5:**
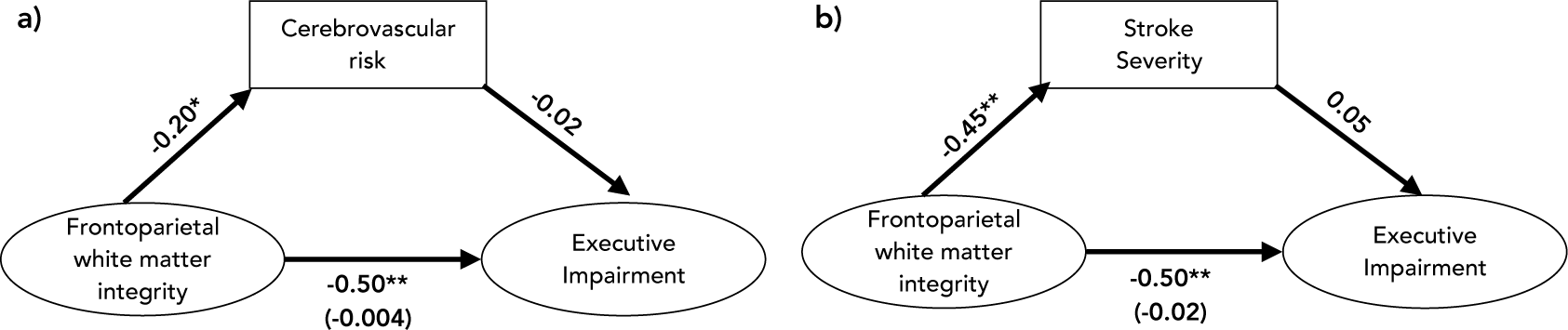
Mediation analysis. Mediation analyses of the relationship between executive impairment and frontoparietal white matter integrity mediated by a) cerebrovascular risk (CVR) and b) stroke severity. *path significant to p<0.05, **p<0.001. Value in brackets indicates the standardized beta estimate of the indirect path.

## Discussion

We demonstrate executive function below what would be expected for age and education level in a cohort of ischemic stroke patients three months after stroke. This is important, as executive dysfunction has been shown to be a significant predictor of quality of life after stroke. A structural equation model was used to assess the complex, multivariate relationship between executive dysfunction and stroke characteristics, cerebrovascular burden and frontoparietal white matter integrity. The location or severity of a stroke on its own cannot predict whether a patient will experience executive dysfunction^11^, which is a significant predictor of quality of life after stroke. Importantly, characteristics of the stroke itself, including measures of functional outcome (mRS), stroke severity (NIHSS) and lesion volume were very poor predictors of executive dysfunction. Our model confirmed that executive dysfunction was associated with the integrity of a key associative tract, the superior longitudinal fasciculus (SLF) within the frontoparietal executive control network.

Nested model-testing confirmed the importance of the integrity of the SLF for predicting executive dysfunction, with significantly worse model fit when SLF integrity was removed from the model. We further interrogated the relationship between executive dysfunction, SLF white matter integrity, cerebrovascular burden and stroke severity, and found the strongest correlation between frontoparietal white matter integrity and executive dysfunction. Mediation analysis revealed that both CVR factors and stroke severity were significant predictors of frontoparietal white matter integrity, confirming the reduced microstructural integrity seen in stroke patients compared to controls. Mediation analysis also confirmed that the significant relationship between executive function and frontoparietal SLF integrity was mediated by cerebrovascular risk and stroke severity. The relationship was no longer significant when controlling for either CVR or stroke severity, with CVR having a stronger mediating effect than stroke severity. This suggests CVR significantly contributes to age-related white matter microstructural degeneration which in turn impacts on executive dysfunction. Cerebrovascular risk factors are potentially modifiable. Our findings are clinically relevant because control of risk factors may reduce degeneration of frontoparietal white matter integrity, which is already burdened by age-related changes. This implies that risk factor management may reduce the burden of executive dysfunction after stroke.

White matter hyperintensities are the most well-known macroscale marker of cerebrovascular burden, but the relationship between WMH and executive function is not clear, and may reflect more global effects on cognition^34^. What is less well appreciated, in relation to executive dysfunction after stroke, is the integrity of ‘normal appearing white matter’ in domain specific cognitive networks. Our model reveals the critical role of microstructural integrity within the SLF which anchors a brain network responsible for executive processing, the frontoparietal network^6^.

The SLF is a major tract in the frontoparietal control network^8,35^ which is specifically vulnerable to the effects of cerebrovascular burden. Ischemic stroke is a clinical manifestation of cumulative vascular risk, but WMHs and brain atrophy from both grey and white matter degeneration occur in the background, with resultant executive dysfunction commonly observed in stroke patients. Our model shows that it may not be the stroke itself that is responsible for executive dysfunction, but rather a multifaceted model that includes the integrity of the major network underlying executive function.

It is important to consider the work in the context of some limitations. We only explored the white matter integrity of the superior longitudinal fasciculus, so we cannot say that degeneration is specific to this tract, and may reflect more global white matter degeneration. We used nested model testing to try to establish the importance of frontoparietal white matter integrity and found a model that included it was indeed a better fit (even when being penalized for increasing model complexity). The SEM was limited to stroke patients, because we were interested in estimating the relationship between stroke severity and executive dysfunction. This made it difficult to control for age-related changes, as age is frequently the biggest predictor of cognitive effects. However, our stroke and control group were age matched, and we showed that executive function was impaired in our stroke group, even when controlling for age and education. We further showed that cerebrovascular risk and white matter hyperintensity burden is increased in stroke patients and white matter integrity of the SLF in reduced in stroke patients compared to age-matched controls. Finally, because stroke characteristics were estimated using a latent variable for the structural equation modelling, we were unable to include a categorical variable representing stroke location. There is some evidence that patients with strokes affecting fronto-subcortical circuits are more likely to develop executive impairment^11^.

## Conclusions

Executive impairment is common after ischemic stroke and is evident even in a relatively mild stroke cohort. Integrity of normal appearing white matter within a large-scale network specific to executive control, predicts executive dysfunction in stroke patients at three months post-stroke. Control of cerebrovascular risk factors may help to mitigate age-related white matter degeneration.

## Data Availability

The data that support the findings of this study are available on reasonable request from the corresponding author. The data are not publicly available as CANVAS is a prospective, live study, with expected completion of data acquisition in mid-2020 for the 5-year scanning timepoint. All requests for raw and analysed data will be reviewed by the CANVAS investigators to determine whether the request is subject to any intellectual property or confidentiality obligations.

## Acknowledgements

We would like to thank our patients and their carers for their time taking part in the experiment over a number of years.

This work was supported by the National Health and Medical Research Council project grant number APP1020526, the Brain Foundation, Wicking Trust, Collie Trust, and Sidney and Fiona Myer Family Foundation.

## Author contributions

Conception and design of study: MV, AB

Acquisition and analysis of data: EW, MK, NE

Drafting a significant proportion of the manuscript: All authors

## Conflicts of interest

The authors have no conflicts of interest to declare.

## Notes

### Competing Interest Statement

The authors have declared no competing interest.

### Clinical Protocols

https://doi.org/10.1111/ijs.12301

